# Distinct cognitive and functional connectivity features from healthy cohorts can identify clinical obsessive-compulsive disorder

**DOI:** 10.1101/2024.09.02.24312960

**Authors:** Luke J. Hearne, B.T. Thomas Yeo, Lachlan Webb, Andrew Zalesky, Paul B. Fitzgerald, Oscar W. Murphy, Ye Tian, Michael Breakspear, Caitlin V. Hall, Sunah Choi, Minah Kim, Jun Soo Kwon, Luca Cocchi

**Affiliations:** QIMR Berghofer Medical Research Institute, Brisbane, QLD, Australia; Centre for Sleep & Cognition & Centre for Translational Magnetic Resonance Research, Yong Loo Lin School of Medicine, Singapore, National University of Singapore, Singapore, Singapore; Department of Electrical and Computer Engineering, National University of Singapore, Singapore, Singapore; N.1 Institute for Health, National University of Singapore, Singapore, Singapore; Integrative Sciences and Engineering Programme (ISEP), National University of Singapore, Singapore, Singapore; Department of Medicine, Human Potential Translational Research Programme & Institute for Digital Medicine (WisDM), Yong Loo Lin School of Medicine, National University of Singapore, Singapore; Martinos Center for Biomedical Imaging, Massachusetts General Hospital, Charlestown, USA; Melbourne Neuropsychiatry Centre, Department of Psychiatry, The University of Melbourne & Melbourne Health, Melbourne, Australia; School of Medicine and Psychology, Australian National University, Canberra, Australia; Central Clinical School, Monash University, Clayton, Australia; Bionics Institute, East Melbourne, Australia; School of Psychological Sciences, College of Engineering Science and Environment, University of Newcastle, Callaghan, Australia; School of Medicine and Public Health, College of Health and Medicine, University of Newcastle, Callaghan, Australia; Program of Neuromodulation, Hunter Medical Research Institute, New Lambton, Australia; Department of Brain and Cognitive Sciences, Seoul National University College of Natural Sciences, Republic of Korea; Department of Neuropsychiatry, Seoul National University Hospital, Republic of Korea; Department of Psychiatry, Seoul National University College of Medicine, Republic of Korea; Faculty of Medicine, School of Biomedical Sciences, University of Queensland, Brisbane, Australia

**Keywords:** OCD, networks, meta matching, big data, transfer learning, fMRI

## Abstract

Improving diagnostic accuracy of obsessive-compulsive disorder (OCD) using models of brain imaging data is a key goal of the field, but this objective is challenging due to the limited size and phenotypic depth of clinical datasets. Leveraging the phenotypic diversity in large non-clinical datasets such as the UK Biobank (UKBB), offers a potential solution to this problem. Nevertheless, it remains unclear whether classification models trained on non-clinical populations will generalise to individuals with clinical OCD. This question is also relevant for the conceptualisation of OCD; specifically, whether the symptomology of OCD exists on a continuum from normal to pathological. Here, we examined a recently published “meta-matching” model trained on functional connectivity data from five large normative datasets (N=45,507) to predict cognitive, health and demographic variables. Specifically, we tested whether this model could classify OCD status in three independent clinical datasets (N=345). We found that the model could identify out-of-sample OCD individuals. Notably, the most predictive functional connectivity features mapped onto known cortico-striatal abnormalities in OCD and correlated with genetic brain expression maps previously implicated in the disorder. Further, the meta-matching model relied upon estimates of cognitive functions, such as cognitive flexibility and inhibition, to successfully predict OCD. These findings suggest that variability in non-clinical brain and behavioural features can discriminate clinical OCD status. These results support a dimensional and transdiagnostic conceptualisation of the brain and behavioural basis of OCD, with implications for research approaches and treatment targets.

## Main

Obsessive-compulsive disorder (OCD) is a disabling mental condition characterised by the presence of intrusive thoughts (obsessions) and/or excessive ritualistic behaviours (compulsions) ^1^. Neurobiological models of OCD supported by genetic, preclinical and neuroimaging studies implicate dysfunction within cortico-striatal-thalamic circuitry ^2–7^. Accurate individual classification, rather than group-level observations, is an important next step in understanding OCD. However, this objective is challenged by the difficulty of acquiring sufficiently large datasets ^8^, which has likely led to over-inflated classification accuracies and poor model generalisability ^9^. Large population-based cohorts like the UK Biobank (UKBB) have been touted as a viable way forward to address the lack of sample size and richness within clinical datasets.

A major challenge in applying “big data” brain models to mental health conditions is the lack of individuals with clinical disorders in such databases. Specifically, in the case of OCD, large publicly available datasets tend not to include individuals with severe or extreme levels of obsessions or compulsions. Nonetheless, contrary to the dominant categorical view of OCD whereby an individual either has or does not have the disorder, several studies support a *dimensional* conceptualisation, with relevant behavioural, neurophysiological, and genetic phenotypes representing a continuum ^10^. This framework suggests that individual brain and behavioural variability associated with subclinical OCD symptoms and dimensional traits (e.g., compulsivity) in the general population may be leveraged to guide research, improve diagnostic tools, and develop targeted personalised treatments for clinical OCD ^11–13^. However, it is uncertain if brain features linked to the expression of subclinical OCD symptoms and traits align with established diagnostic (e.g., DSM-V) and neurophysiological (e.g., cortical-striatal-thalamic loops) models of the disorder. Accordingly, the success of models based on “big data” relies on whether obsessive-compulsive features do indeed form a continuum from healthy individuals to clinical OCD with overlapping brain characteristics.

A recent approach, termed “meta-matching”, demonstrated successful resting-state functional connectivity (RSFC) brain-behaviour model generalisation from large to small datasets in healthy cohorts ^14,15^. Meta-matching leverages the observation that many cognitive, health and demographic variables are correlated. Thus, brain connectivity patterns useful for classifying a known variable (e.g., age) are also likely to be useful in classifying an unseen, *correlated* variable in a different dataset (e.g., memory performance). This approach demonstrated impressive classification performances, even in small to moderate sample sizes ^15^, suggesting it may be useful to inform clinical neuroimaging studies.

In the current work, we started by assessing whether a meta-matching model, trained on five large healthy datasets, was useful in identifying persons with an OCD diagnosis. We then investigated the brain features generated by the model and, in line with the cortical-striatal-thalamic loop model of OCD, hypothesised a large contribution of RSFC features associated with cortico-striatal circuits ^3,16^. We also explored the behavioural and physiological phenotypes derived from the large healthy datasets contributing to OCD prediction. Finally, to further evaluate the biological validity of the meta-matching model, we assessed the relationship between its connectivity features and genetic expression maps previously associated with compulsivity in the general population ^17^.

## Results

First, we established whether a brain-based classifier trained on healthy individuals could accurately identify persons with an OCD diagnosis. To achieve this, we used a meta-matching model ^14^ that had been trained to predict 458 variables reflecting different aspects of physical and mental health, cognition, and behaviour. This model used whole-brain RSFC calculated across five source datasets, encompassing a total of 45,507 participants (**Methods**). Our local, to-be-classified dataset was a clinical OCD sample and matched healthy controls from three independent sites (N=345, n_OCD_ = 199, n_HC_ = 146). We refer to the data used in the meta-matching model as “normative” to distinguish it from the local healthy control data. Model accuracy was assessed in held-out test samples in a standard five-fold cross-validation scheme (**Methods**).

### Normative meta-matching RSFC brain-behaviour models can be used to classify OCD

We found the meta-matching model could accurately classify unseen RSFC data as OCD or healthy controls (median balanced accuracy = 61.1%, *p* = 0.003, **Figure 1A**). Brain features utilised in the meta-matching model are shown in **Figure 1B** (**Methods**). Positive brain feature weights indicate that higher RSFC was observed in the OCD group for a given brain region compared to the local dataset of healthy controls or vice versa. For example, visual cortex regions demonstrated high feature weights and, therefore, higher RSFC in OCD.

**Figure 1.**
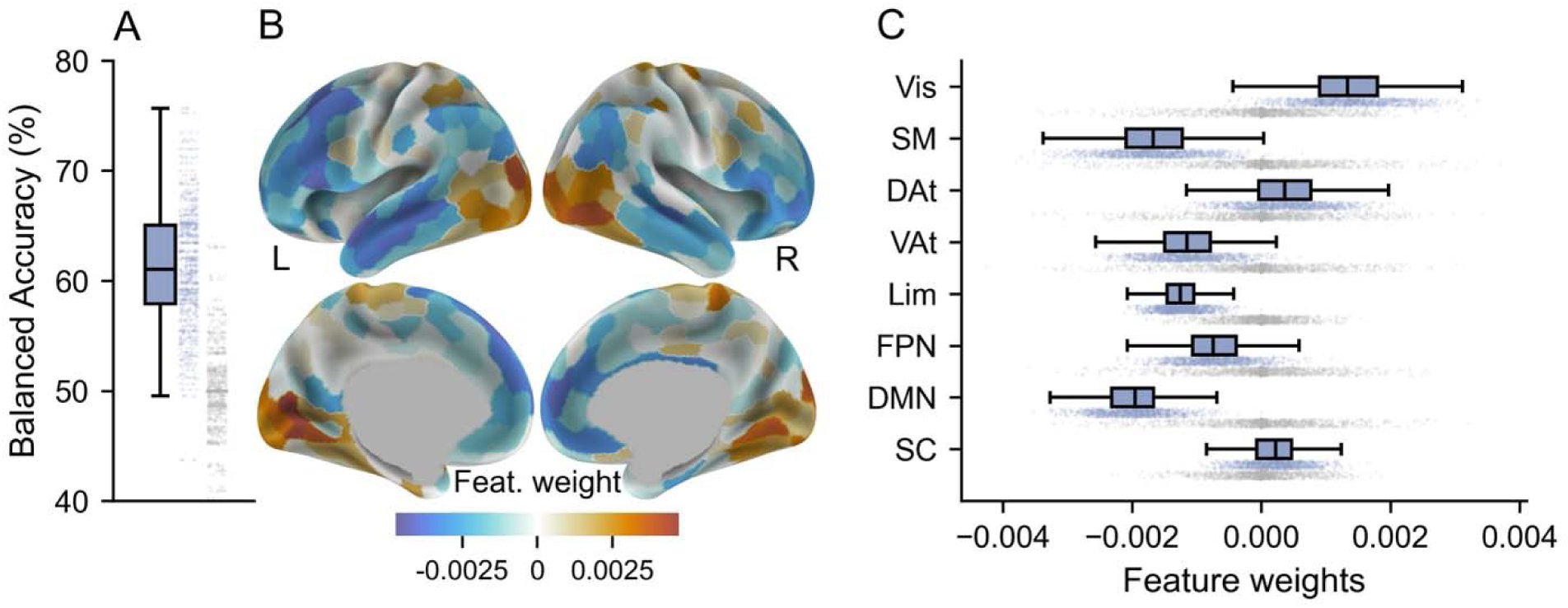
Meta-matching model performance and brain features. **A.** Balanced accuracy (y-axis) classification performance in out-of-sample data for the meta-matching model (blue) and shuffled permutations (grey). Individual data points represent the variability across the cross-validation folds (200 iterations of 5 folds). **B.** Visualisation of the averaged brain features used in the model. **C.** Brain features averaged into canonical functional networks. Brain networks: FPN; control/frontoparietal, DMN; default-mode, DAt; dorsal attention, Lim; limbic, VAt; salience/ventral attention, SM; somatomotor, SC; subcortical, Vis=visual). Boxplots: centre line; median; box limits; upper and lower quartiles; whiskers; 1.5x interquartile range.

Next, we averaged the regional weights into eight canonical functional brain networks ^18^. All networks but the subcortical and limbic networks showed significant contributions to the successful predictions (**Figure 1C**, **Supplementary Table 1**). Visual and dorsal attention networks were associated with increased OCD functional connectivity, whereas all other significant networks were associated with decreased connectivity in OCD (**Figure 1C**).

### Evaluating the predictive contribution of cortico-striatal circuits

OCD has been consistently associated with abnormal cortico-striatal circuit function ^2,6,16,19^. Thus, we tested whether such circuits were more predictive of OCD status than other equally sized sets of subcortical-cortical functional brain connections. An independent dataset (N=250) was used to map the cortico-striatal circuits of interest ^20^ (**Methods**). As expected, connectivity patterns involving the striatal seed regions (NAcc; Nucleus Accumbens, dCaud; dorsal Caudate, dPut; dorsal Putamen, vPut; ventral Putamen; **Supplementary Figure 1**) highlighted a largely striatal-frontal pattern of connectivity (**Figure 2A**). Using this data, “circuits of interest” were defined as the most highly connected cortical regions with positive RSFC (10 regions per hemisphere, per seed, cortical areas within black contours in **Figure 2A**).

**Figure 2.**
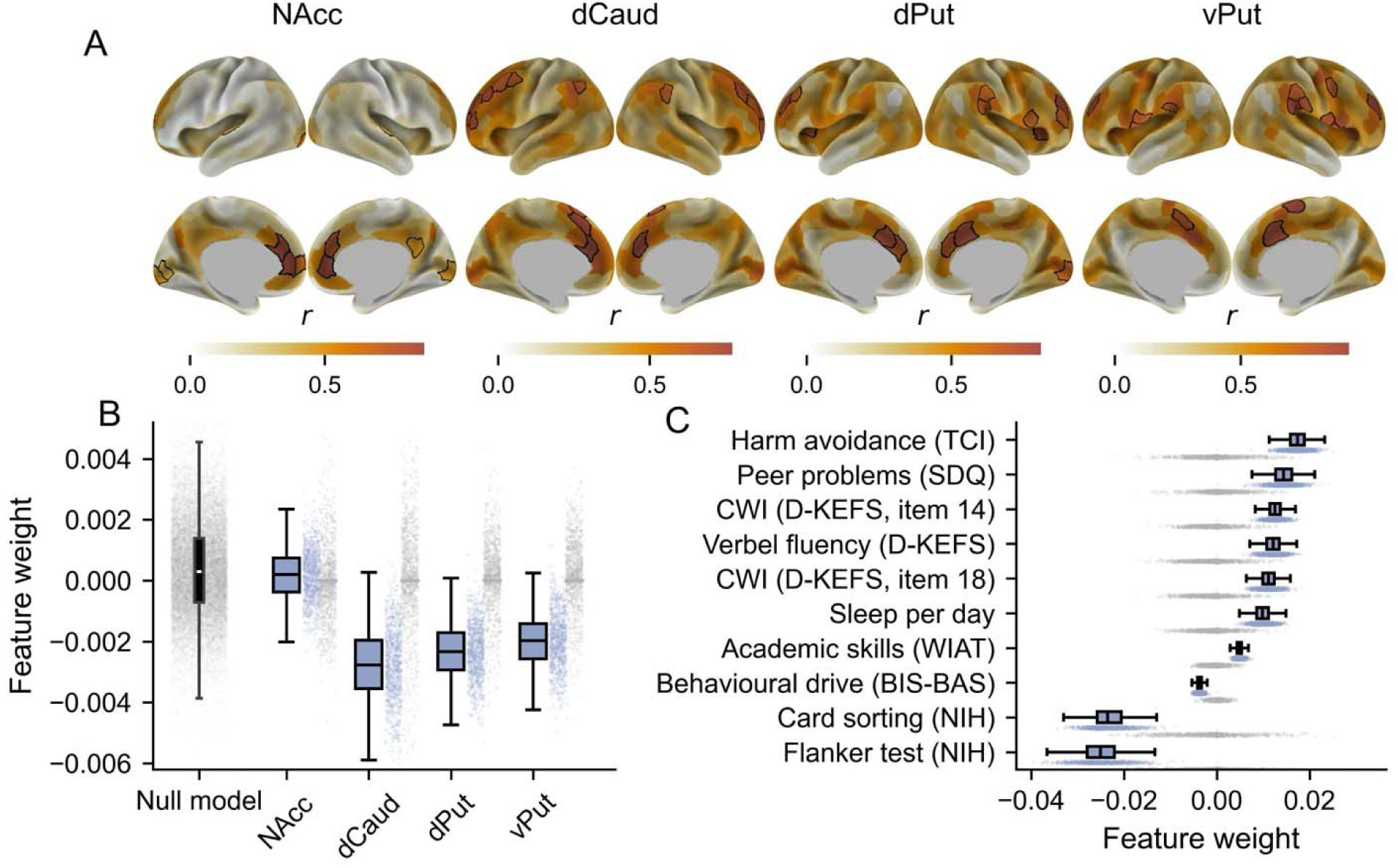
Predictive weights within cortico-striatal circuits. **A.** Group- and hemisphere-averaged functional connectivity maps from 250 unrelated HCP participants when seeding each subcortical region of interest. Black borders indicate the cortico-striatal circuits of interest based on cortical regions with the highest positive connectivity values. NAcc; Nucelus Accumbens, dCaud; dorsal Caudate, dPut; dorsal Putamen, vPut; ventral Putamen (Supplementary Figure 1). **B.** Comparison between a null model of random subcortical-cortical brain feature weights (grey, far left) and the circuits of interest for the meta-matching model (blue). Circuit-specific shuffled permutations are also shown in grey. Individual data points represent the variability across the cross-validation folds (200 iterations of 5 folds). **C.** Top ten meta-matching phenotypes with the largest feature weights. Positive values indicate the OCD cohort had higher weights than the local cohort healthy controls and vice versa (e.g., lower functional connectivity in the dCaud pathway in panel **B**, or increased harm avoidance in panel **C**). TCI; Temperament and Character Inventory, SDQ; Strengths and Difficulties Questionnaire, CWI; Colour Word Interference, D-KEFS; Delis-Kaplan Executive Function System, WIAT; Wechsler Individual Achievement Test, BIS-BAS; Behavioural Inhibition and Behavioural Activation Systems, NIH refers to the NIH toolbox. Boxplots: centre line; median; box limits; upper and lower quartiles; whiskers; 1.5x interquartile range.

We observed consistent outlier brain features in the dorsal caudate, dorsal putamen, and ventral putamen circuits compared to shuffled permutations (dCaud; *p*= 0.01, dPut; *p* = 0.01, vPut; *p* = 0.01; **Figure 2B**). The dorsal caudate and dorsal putamen circuits also had larger negative feature weights than would be expected from any random set of subcortical-cortical brain connections (dCaud; *p* = 0.02, dPut; *p* = 0.04, **Figure 2B** and **Supplementary Table 2**). Note that, as before, the sign indicates the direction of the effect. For example, connectivity is lower in OCD in the dorsal putamen pathway compared to the local dataset healthy controls (**Figure 2B**).

### Trained phenotypes contributing to OCD predictions

We next sought to establish which phenotypes from the normative population data used in the meta-matching model were useful for classifying OCD. Specifically, the meta-matching approach transforms brain connectivity data into 458 variables representing different health, cognitive and behavioural phenotypes derived from several datasets. We investigated which of these variables were useful in predicting OCD status. Of those 458 variables, only two were significantly related to OCD predictions: (i) the Dimensional Change Card Sort Test, a measure of cognitive flexibility; and (ii) the Flanker Inhibitory Control and Attention test (*p*_FDR_ < 0.05, **Supplementary Table 3**).

Other variables that were highly weighted (e.g., *p*_FDR_ < 0.06) included cognitive measures such as performance on the Colour Word Interference test (a test of cognitive flexibility), mental health (e.g., excessive worrying indexed by harm avoidance), and behavioural variables (e.g., increased sleep per day) (**Figure 2C**).

### Relationship between meta-matching model brain features and genes implicated in OCD

Next, we sought to understand the link between the brain connectivity features used in the meta-matching model and the known role of genetics in the development of OCD ^21,22^. A recent GWAS meta-analysis identified that variability within the KIT, GRID2, ADCK1, and WDR7 genes are associated with an increased likelihood of compulsive symptoms ^17^. Broadly speaking, the KIT gene is involved in cell proliferation and survival, while the GRID2 gene supports glutamate signalling in cerebellar Purkinje cells ^23,24^. The ADCK1 gene maintains mitochondrial function, and the WDR7 gene aids proteins involved in neurotransmission ^25,26^. To investigate these gene expression in the brain for these specific genes, we utilised preprocessed regional microarray expression data provided by the Allen Human Brain Atlas via the Abagen toolbox ^27,28^. Brain-wide regional gene expression was correlated with connectivity features generated by the meta-matching model (cortical features shown in **Figure 1B**).

Three of the regional gene expression maps could be reliably linked to the brain connectivity features in the meta-matching model (GRID2, *r* = -0.31, *p* <0.001; WDR7, *r* = -0.30, *p* <0.001; ADCK1, *r* = -0.14, *p* = 0.03) (**Figure 3**). When considering the subcortex, only WDR7 demonstrated a significant correlation (WDR7, *r* = -0.64, *p* = 0.01, see **Supplementary Table 4**). For each significant relationship, higher regional gene expression was associated with negative connectivity features (i.e., regions with lower RSFC in OCD compared to the local dataset healthy control cohort). Note that statistical significance was declared using the spin test ^29^, followed by FDR correction for multiple comparisons.

**Figure 3.**
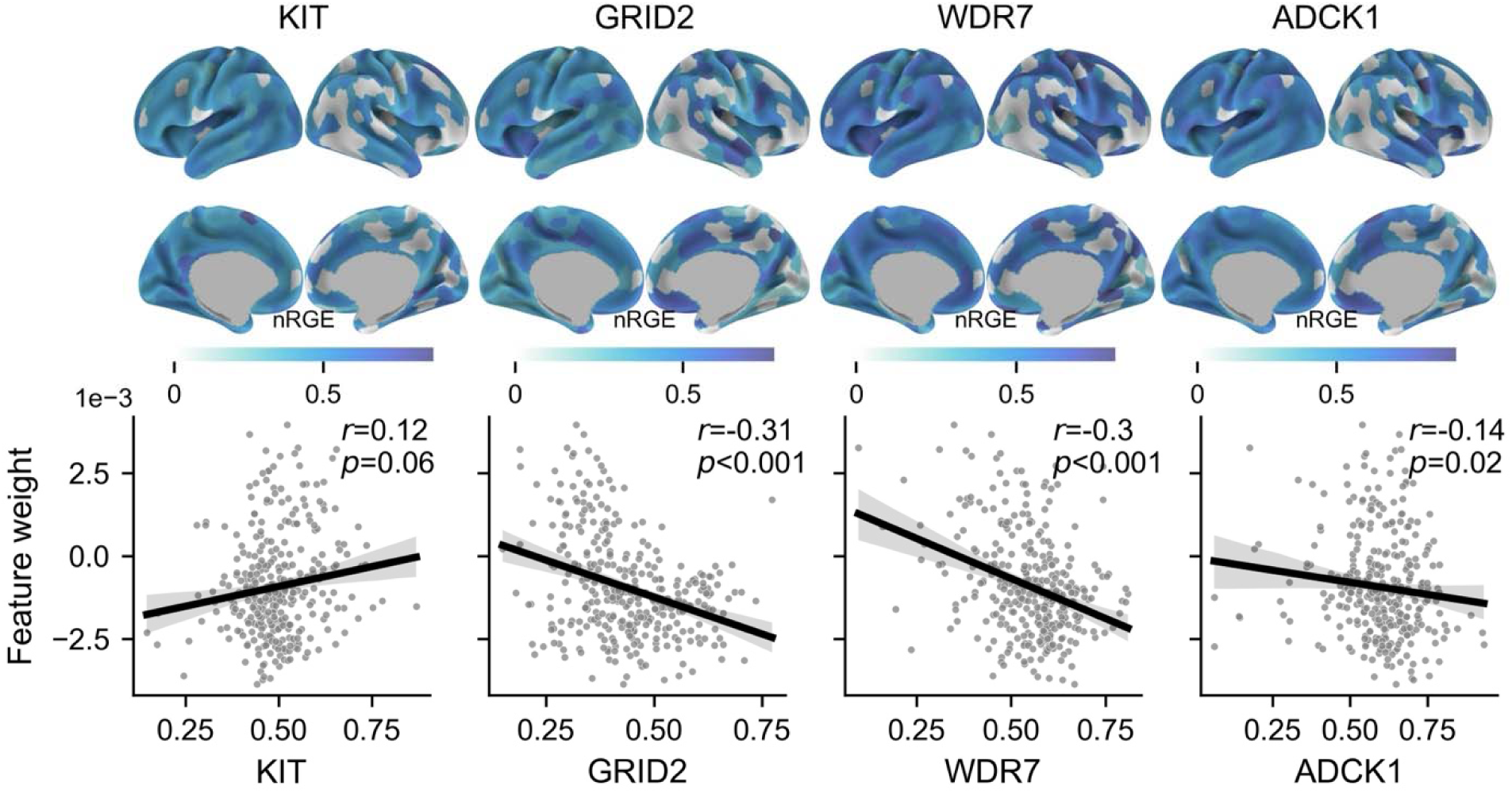
Regional gene expression associated with meta-matching predictive feature weights. The top panel shows the normalised regional gene expression (nRGE) for each of the four genes of interest. The bottom panel displays the linear relationship between each nRGE cortical map and the meta-matching model connectivity feature weights.

### Control analyses

We contrasted the current results obtained using multilayer meta-matching ^14^ with the original meta-matching model ^15^. The key difference between these models is the increased number of training datasets and phenotypes in the multilayer meta-matching (the original model only used data from the UKBB for training). The multilayer meta-matching performance was not statistically different from the original model (*p* = 0.44, **Supplementary Figure 2**).

We also performed a control experiment using a logistic regression model to classify OCD patients (**Methods**). Classification performance for this model was not statistically different to the meta-matching models (*p* = 0.53, **Supplementary Figure 2**). Likewise, the brain features extracted from the models were significantly correlated (cortex: *r* = 0.92, *p* < 0.001, subcortex: *r* = 0.82, *p* < 0.001).

Finally, when developing brain-behaviour prediction models, it is common practice to attempt to remove (*deconfound)* covariates that may bias the results (e.g., age, sex, site, head motion). Accordingly, we have presented results adjusted for site, age and sex (see **Methods**). Classification performance was not statistically different between the deconfounded and non-deconfounded models (*p* = 0.93), suggesting that variables such as age, sex, and head motion had a limited impact on the results (**Supplementary Figure 2**).

## Discussion

In this study, we tested if a brain-behaviour model trained on large, relatively “healthy” (i.e., non-clincal) datasets could classify individuals with clinical OCD. Despite the lack of individuals with a clinical diagnosis of OCD and relevant measurements (e.g., symptom scales) in the training data, the model demonstrated above-chance performance. This finding supports the conceptualisation of OCD as a continuum comprising core behavioural, neurophysiological, and genetic aspects ^30^. Specifically, the predictive power of cognitive phenotypes developed in healthy training data - cognitive flexibility, attention, and inhibitory control - highlights the value of these non-specific transdiagnostic measures for understanding OCD ^31^. Likewise, the results confirm the key role of dysregulated frontostriatal functional connectivity in OCD ^3,7^ and their relevance in the expression of subclinical phenotypes. Finally, we were able to establish a link between OCD-related brain connectivity features and gene expression associated with compulsiveness in the general population ^10,17^.

The ubiquitous nature of the brain-behaviour mappings used by the meta-matching model is consistent with the idea that OCD pathology is supported by mental processes and behaviours that cut across traditional diagnostic categories and exist on a spectrum between healthy and diseased states ^12,32,33^. Accordingly, cognitive deficits like the one indexed by the Dimensional Card Sorting task or the Flanker Task are believed to be markers of general psychopathology detectable across various neurological and psychiatric conditions ^34,35^. In addition to the above cognitive processes, our findings highlight the strong predictive weight of harm avoidance or escape “coping”, which are more specific to OCD. Excessive harm avoidance and its underpinning processes are indeed thought to be a core mechanism of compulsive behaviour ^36,37^. These findings suggest that the subclinical “healthy population” variability in core cognitive processes is useful for classifying clinical OCD and may contribute to the emergence of OCD-specific behaviours.

Our analysis of the meta-matching model’s brain connectivity feature weights supports the hypothesis that abnormal RSFC within cortico-striatal circuits represents a hallmark of OCD pathophysiology ^16,19^. Specifically, resting-state connectivity of the dorsal caudate and putamen circuits was one of the largest predictors across all possible subcortical-cortical brain connections. These circuits have been previously associated with cognitive deficits and sensorimotor symptoms observed in OCD, such as planning, decision-making, and repetitive behaviours ^38–42^. Therefore, our findings provide additional motivation to study the neural mechanisms underpinning abnormalities in frontostriatal activity in people with clinical OCD. Investigating the brain basis of changes in frontostriatal circuit activity is key to improving the efficacy of invasive neurosurgical treatments ^43^ and the development of mechanism-sensitive non-invasive therapies like transcranial magnetic stimulation ^44^.

Similar to prior work attempting to use neuroimaging data to classify OCD individuals, we also observed that, compared to other macroscale networks, the default-mode and sensory networks contained the largest predictive features ^45,46^. The default mode network, which has been linked to internal monitoring ^47^, may align with ruminations and excessive self-monitoring observed in OCD ^48^. Likewise, sensorimotor systems have been associated with inhibition problems at the core of several OCD symptoms ^49^. Supporting the importance of functional brain networks in OCD, the expression patterns of genes previously associated with clinical OCD and subclinical compulsivity ^17^ correlated with brain-wide predictive feature patterns. Specifically, brain regions with lower RSFC in OCD, relative to controls, tended to exhibit higher gene expression. This suggests that differences in the regional brain expression of genes broadly associated with neural function and communication (e.g., GRID2 cerebellar glutamate signalling, ADCK1 mitochondrial function, WDR7 neurotransmission) can be tied to brain connectivity in OCD. Collectively, our findings underscore the importance of the neurobiological features leveraged by the meta-matching model to explain and predict clinical OCD.

Our results also shed light on the broader feasibility of transfer learning from general population datasets to clinical populations ^50,51^. Unlike prior studies, we did not observe a boost in classification performance due to meta-matching when compared to a simple regression model ^14,15,52^. There are likely multiple reasons for this discrepancy, the most obvious being that some critical aspects of variability associated with clinical OCD are not observable in brain-behaviour mappings trained in a healthy population. Thus, a key future research direction is improving meta-matching performance to a clinically useful level, including diagnosing OCD from brain connectivity measures.

In conclusion, we have demonstrated that large healthy normative imaging datasets can be used to classify and advance knowledge of the brain and behavioural bases of clinical OCD. Our investigation into cortico-striatal circuits, functional networks, and OCD-specific brain-wide gene expression patterns, provides strong evidence for the biological relevance of the adopted meta-matching model. These results support the study of OCD via a transdiagnostic framework defined by core neuro-behavioural constructs ^32^. The findings also motivate further investigations into using meta-matching and transfer learning to improve the biologically grounded classification of psychiatric conditions beyond OCD.

## Methods

### Datasets

Data used in the current study were pooled across three independent datasets collected in Brisbane ^44^, Melbourne (clinical trial registration ACTRN12619000008123), and Seoul ^53^. The specifics of each dataset are described in the following section. The final sample size after the exclusion of people with excessive head motion (below) was N = 345 (N_OCD_ = 199, N_HC_ = 146). Demographics and clinical characteristics for each of these datasets are described in **Table 1**. In addition, 250 unrelated participants from the Human Connectome Project ^54^ were used to map cortico-striatal circuits of interest. The relevant local ethics committees approved each study. Likewise, all studies complied with the ethical standards of the relevant national and institutional committees on human experimentation.

**Table 1.**
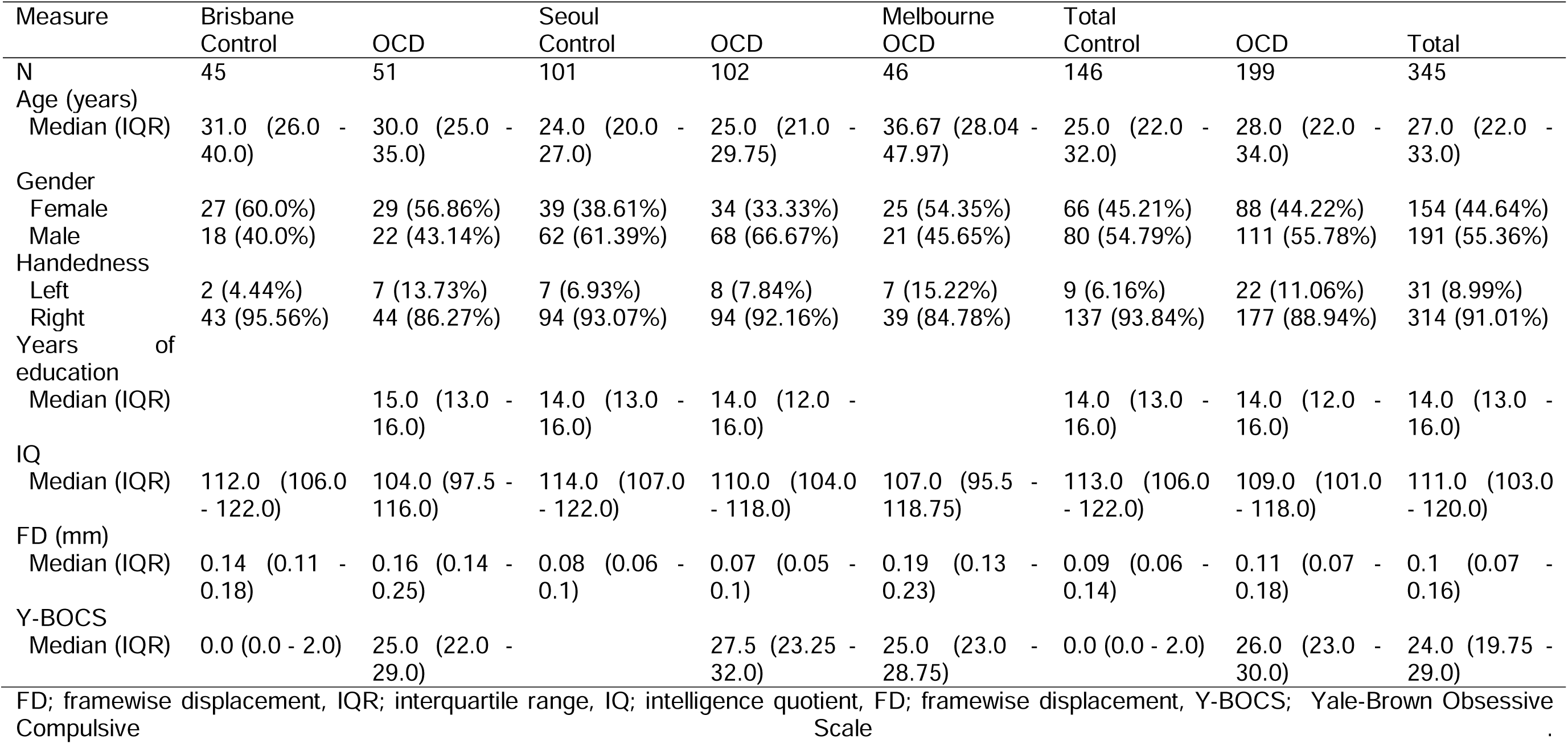
Sample demographics.

#### Brisbane

Fifty-eight adult participants with a clinical diagnosis of OCD and 45 controls were recruited across Australia as part of a registered randomised-controlled clinical trial (ACTRN12616001687482). In total, seven participants from the OCD cohort were excluded due to excessive head motion, anatomical abnormalities and missing and/or corrupted data (see brain imaging preprocessing section below). Details regarding participant recruitment and inclusion criteria have been reported elsewhere ^44,55^.

#### Melbourne

Forty-six adult participants with a clinical diagnosis of OCD were recruited across Australia as part of a registered randomised-controlled clinical trial (ACTRN12619000008123). The trial aimed to recruit a total of 75 participants, but it was prematurely terminated because of the COVID-19 pandemic. The study was approved by the Alfred Health Research Ethics Committee (Melbourne, Australia). Written informed consent was obtained from all participants. Details regarding participant recruitment and inclusion criteria are reported in the supplementary material.

#### Seoul

102 medication-free adult participants with a clinical diagnosis of OCD and 101 controls were recruited as part of a previous study ^53^. The Institutional Review Board of Seoul National University Hospital approved the study. Written informed consent was obtained from all participants (any minors who participated in the study required consent from the individual and their caretakers). Details regarding participant recruitment and inclusion criteria have been reported elsewhere ^53^.

### Brain imaging data acquisition

#### Brisbane

Brain imaging data were acquired on a 3T Siemens Prisma MR scanner equipped with a 64-channel head coil at the Herston Imaging Research Facility, Brisbane, Australia. Whole brain echo-planar images were acquired with the following parameters: voxel size = 2 mm^3^, TR = 810 ms, multiband acceleration factor = 8, TE = 30 ms, flip angle = 53°, field of view = 212 mm, 72 slices. The resting state acquisition was about 12 minutes in length (880 volumes). Structural brain images were acquired with the following parameters: voxel size = 1 mm^3^, TR = 1900 ms, TE = 2.98 ms, 256 slices, flip angle = 9°. Anterior-to-posterior and posterior-to-anterior spin echo fieldmaps were also acquired.

#### Melbourne

Brain imaging data were acquired on a 3T Siemens Prisma MR scanner equipped with a 64-channel head coil at The Royal Melbourne Hospital, Melbourne, Australia. The functional and structural brain imaging sequences were identical to the Brisbane site.

#### Seoul

Brain imaging data were acquired on a 3T Siemens Trio MR scanner equipped with a 12-channel head coil at the Seoul National University Hospital. Whole brain echo-planar images were acquired with the following parameters: voxel size = 1.9 mm x 1.9 mm x 3.5 mm, TR = 3500 ms, TE = 30 ms, flip angle = 90°, field of view = 240 mm, 35 slices. The resting state acquisition was about 7 minutes in length (116 volumes). Structural brain images used in the preprocessing pipeline were acquired with the following parameters: voxel size = 1 mm x 0.98 mm x 0.98 mm, TR = 1670 ms, TE = 1.89 ms, 208 slices, flip angle = 9°.

### Brain imaging data processing

All brain imaging data were preprocessed using fMRIprep (version 23.2.0) ^56^. Briefly, the data were skull stripped, corrected for susceptibility distortions (multiband data only), coregistered to the anatomical image and slice time corrected. The data were then resampled to a standard template space (HCP CIFTI surface and volume) (see supplementary for full details). The data were then downsampled into region-specific timeseries using the Schaefer 400 brain parcellation ^57^, with an additional 19 subcortical and cerebellar regions ^58^. This specific atlas was required to use the previously published meta-matching model ^15^.

The resulting timeseries were denoised via Nilearn using a standard, previously benchmarked pipeline ^59^. For the main results, we employed a denoising strategy that regressed 24 motion parameters (six motion parameters, their temporal derivatives and quadratics of all regressors), average white matter signal, cerebrospinal fluid signal, global signal, as well as cosine transformation basis regressors. Framewise displacement was used to identify participants with large amounts of head motion. Specifically, any participant with less than 5 minutes of data with a framewise displacement of less than 0.5 mm was excluded from further analysis (N = 2 from the OCD cohort at the Brisbane site).

Finally, consistent with the original meta-matching model, Pearson correlation was conducted on the denoised timeseries to estimate RSFC. Thus, for each participant, there was a single 419 x 419 symmetric RSFC matrix. The unique, lower triangle values from these matrices were indexed, resulting in a final array of RSFC values by participants (87,543 x N), which were used as features in the classification models.

### Meta-matching model

To perform meta-matching, we used the openly available multilayer meta-matching model published by Chen et al. (2022) (V2.0; https://github.com/ThomasYeoLab/Meta_matching_models). The model was trained on five source datasets: the UKBB ^N = 36,834;, 60^, the Adolescent Brain Cognitive Development study ^ABCD, N = 5,985;, 61^, the Healthy Brain Network project ^HBN, N = 930;, 62^, the enhanced Nathan Kline Institute-Rockland sample ^eNKI-RS, N = 896;, 63^, and the Genomics Superstruct Project ^GSP, N = 862;, 64^. Crucially, the samples used in these studies reflect individuals who are healthier than the general population ^65^. The UKBB did include some individuals who self-reported a lifetime OCD diagnoses, regardless of whether they were exhibiting symptoms at the time of imaging. However, given their small number (approximately 0.06%)^66^, it is unlikely that this sub-sample could drive our results. Age varied across these datasets, ranging from children, adolescents, younger adults, and older adults (see Chen et al., 2023 for further details).

This meta-matching model uses a combination of a fully connected feedforward deep neural network and multiple kernel ridge regression models to predict 458 *non-brain imaging phenotypes* from RSFC (87,543 edges). The 458 phenotypes were derived from the source datasets, representing different aspects of physical and mental health, cognition, and behaviour. Comprehensive details regarding the model and its training data are available elsewhere ^14,15^. In brief, the 458 predictions are made by two parallel approaches. The first uses kernel ridge regression models to generate predictions from RSFC for each dataset, resulting in 229 phenotypic predictions (UKBB=67, ABCD=36, HBN=42, eNKI-RS=61, GSP=23). The second uses RSFC to predict UKBB phenotypes (67) via a fully connected deep neural network and then uses the outcomes of this first layer as input to the remaining kernel ridge regression models (ABCD=36, HBN=42, eNKI-RS=61, GSP=23).

The 458 phenotypes predicted by the meta-matching model are not specific to our research problem: predicting OCD diagnosis. Thus, we employed a procedure called “stacking”, whereby a final regression model is used to predict the phenotype of interest. Specifically, the 458 predicted phenotypes were used in a logistic regression to predict OCD status. This was implemented using a regularised logistic regression in sklearn (*LogisticRegressionCV*) with the *liblinear* solver where the best *C* hyperparameter was selected via a nested five-fold cross-validation ^67^.

### Cross-validation and model performance

A five-fold cross-validation scheme was employed to assess the accuracy of the predictions. The cross-validation was repeated 200 times to generate a distribution of classification performance measures. Within the training set, site harmonisation was performed using ComBat with OCD status as a covariate of interest ^68,69^, a validated method for correcting multi-site data. Likewise, linear regression was used to remove variability associated with age and gender. These deconfounding models were applied separately to the test data within each cross-validation fold. A statistical analysis of the success of this deconfounding procedure is described below (**Control Analyses**). Model accuracy for each cross-validation iteration was assessed using balanced accuracy.

As in prior work ^70^, we evaluated the performance compared to chance via permutation testing. Specifically, each model was rerun after shuffling the RSFC data, effectively severing the link between RSFC and OCD status. This was repeated 1000 times, generating a null distribution of performance classifications for each model. The null distribution was compared to the real classification by calculating the percentile of the average classification value compared to the null distribution.

### Examination of brain connectivity features

To assess the importance of each functional connectivity edge in predicting OCD status, the Haufe transform ^71^ was used to calculate feature weights. We used these weights to assess the importance of functional brain networks (**Figure 1**) and cortico-striatal circuits (**Figure 2**), as well as the relationship between model features and gene expression (**Figure 3**).

#### Cortico-striatal brain circuits

We mapped four key cortico-striatal circuits by seeding the Nucleus Accumbens (NAcc), dorsal Caudate (dCaud), dorsal Putamen (dPut), and ventral Putamen (vPut) in an independent dataset (N=250 from the Human Connectome Project, HCP; see **Supplementary Material**).

The brain connectivity feature weights of these distinct brain circuits were examined and compared to two null models. The first compared the brain connectivity weights within the circuits to the identical weights from permuted models (described above). The second approach compared the circuit weights to similarly sized sets of connections drawn from a distribution of random subcortical-cortical empirical weights. The null distributions were compared to the real classification by calculating the percentile of the mean real classification value compared to the null distribution.

#### Region-wise brain connectivity features, functional brain networks, and gene expression

Brain connectivity feature weights were averaged across cross-validation folds for each brain region in the adopted brain parcellation. We then performed two analyses on these region-wise values. First, these values were further averaged into eight functional brain networks (control/ frontoparietal, default-mode network, dorsal attention, limbic, salience / ventral attention, somatomotor, subcortical and visual) ^18^ and compared to null permutations. Second, we tested whether brain-wide gene expression in four genes highlighted by a recent GWAS meta-analysis of subclinical compulsive symptoms (KIT, GRID2, ADCK1, WDR7) ^17^ were associated with brain connectivity features. Specifically, regional microarray expression data were collected from six post-mortem brains (one female, ages 24 to 57, average age 42.5 ± 13.38 years) provided by the Allen Human Brain Atlas. The data were processed using the Abagen toolbox in MNI space ^27,28^. When comparing cortical maps, we employed the spin test ^29^. For subcortical maps, we utilised a standard permutation shuffling approach (10,000 permutations in both scenarios) ^72^.

#### Meta-matching phenotype feature weights

Similar to the Haufe transform between predictions and RSFC inputs described above; an analogous analysis was conducted to examine the predictive value of the 458 training dataset phenotypes in the meta-matching model. In this context, a positive weight indicates that the meta-matching model predicted a higher score for a given phenotypic variable for the local dataset OCD patients than the healthy controls, and vice versa. To give a concrete example, the phenotype neuroticism was measured in the UKBB (one of the training normative datasets), however this was not measured in the local OCD and healthy control datasets. If this phenotype feature has a high weight, the meta-matching model has predicted that OCD patients would report higher scores on the neuroticism scale than the local dataset healthy controls. Thus, in this scenario, we would conclude that the RSFC from OCD patients in the local data is similar to RSFC in the UKBB that reported high scores on the neuroticism scale. As in the brain feature connectivity weights, the average empirical value was compared to shuffled null permutations and subsequently FDR corrected (N_components_ = 458).

### Control analyses

#### Alternative model comparisons

The meta-matching model was compared to a logistic regression model in which all RSFC values were used to predict OCD status. Aside from the shape of the input data, this model was identical to the “stacking” logistic regression described above. This baseline model represents a standard comparison model that might be used to predict OCD diagnosis.

We also contrasted the meta-matching model with a prior version (V1.1), which only included a single training dataset (the UKBB, He et al., 2022). Models were compared to each other via the corrected resample *t*-test as a standard *t*-test would not be valid ^73,74^.

#### Deconfounding

Predictive models can capture confounding effects correlated with the outcome rather than the brain features of interest (e.g., age, site, head motion) ^75–77^. To explore this, we compared model performance before and after deconfounding using the corrected resample t-test.

#### Impact of global signal regression

Denoising fMRI data from non-neuronal sources is a crucial preprocessing step in RSFC analyses ^59^. The training data used in the meta-matching model utilised a variety of denoising approaches with and without global signal regression. For completeness, we compared model performance with and without global signal regression in the local dataset using the corrected resample t-test (*p* = 0.41, see **Supplementary Figure 2**).

## Data and code availability

Code used to generate the results are available on GitHub (https://github.com/ljhearne/CBN_MetaMatch_public). The version of the meta-matching model used in the current work is available online (V2.0; https://github.com/ThomasYeoLab/Meta_matching_models). De-identified participant data for research purposes are available on request for data collected at the Brisbane^44^ and Melbourne sites. De-identified participant data for research purposes are available on request for data collected at the Seoul site from the original authors^53^.

## Acknowledgements

This work was supported by the Australian NHMRC (GN2001283 and GNT2027597, L.C). A.Z. and L.J.H were supported by research fellowships from the NHMRC (APP1118153, APP1194070, respectively). PBF is supported by a National Health and Medical Research Council of Australia Investigator grant (1193596). BTTY is supported by the NUS Yong Loo Lin School of Medicine (NUHSRO/2020/124/TMR/LOA), the Singapore National Medical Research Council (NMRC) LCG (OFLCG19May-0035), NMRC CTG-IIT (CTGIIT23jan-0001), NMRC STaR (STaR20nov-0003), Singapore Ministry of Health (MOH) Centre Grant (CG21APR1009), the Temasek Foundation (TF2223-IMH-01), and the United States National Institutes of Health (R01MH120080 & R01MH133334).

