## Supplemental Information for "Distinct cognitive and functional connectivity features from healthy cohorts can identify clinical obsessive-compulsive disorder"

### Supplementary Text

#### Melbourne dataset

Data used in the current study were from a brain stimulation trial conducted in Melbourne. The trial was conducted at one academic centre (Epworth Centre for Innovation in Mental Health, Epworth Healthcare, Australia). People with a primary diagnosis of OCD (DSM-V criteria) for at least 12 months with moderate to severe symptoms (total Y-BOCS score greater than 16) were recruited across Australia. Inclusion criteria were 18-65 years of age, capacity to give informed consent, stable pharmaceutical treatment in the past three months, and ability to attend 15 consecutive weekday brain stimulation sessions. Exclusion criteria were: Y-BOCS below 17, pregnancy (or intention to fall pregnant), high risk of suicide attempt determined by the clinical PI, psychotic disorder, diagnosis of bipolar I or II disorders, Tourette’s syndrome, history of seizures or neurological disorders, history of substance abuse in the past six months, and contraindications to MRI or TMS. Participants were withdrawn from the trial if they missed more than two consecutive brain stimulation sessions or four sessions in total, or if they changed psychotropic medication (no participant was excluded due to this criteria). 49 participants were enrolled in the trial, with 46 completing the required baseline data collection used in the current study. Of the three withdrawals, two became uncontactable after enrolment, and one withdrew during the baseline data collection process (no MRI was collected).

#### Brain imaging preprocessing

Results included in this manuscript come from preprocessing performed using fMRIPrep 23.2.0 ^1,2^, which is based on Nipype 1.8.6 ^3,4^. Below, we have inserted the fMRIprep-generated methods with minimal adjustments to reflect differences across the three local datasets.

*Preprocessing of B0 inhomogeneity mappings*

In the datasets with multiband functional acquisitions (Brisbane, Melbourne). A B0-nonuniformity map (or fieldmap) was estimated based on two (or more) echo-planar imaging (EPI) references with topup ^5^.

*Anatomical data preprocessing*

The T1w image was corrected for intensity non-uniformity (INU) with N4BiasFieldCorrection ^6^, distributed with ANTs 2.5.0 ^7^. The T1w-reference was then skull-stripped with a Nipype implementation of the antsBrainExtraction.sh workflow (from ANTs), using OASIS30ANTs as target template. Brain tissue segmentation of cerebrospinal fluid (CSF), white-matter (WM) and gray-matter (GM) was performed on the brain-extracted T1w using fast ^FSL 5.0.9, 8^. For the Melbourne dataset, which had two T1w images, an anatomical T1w-reference map was computed after registration of two T1w images (after INU-correction) using mri_robust_template ^FreeSurfer 7.3.2, 9^. Brain surfaces were reconstructed using recon-all ^10^, and the brain mask estimated previously was refined with a custom variation of the method to reconcile ANTs-derived and FreeSurfer-derived segmentations of the cortical gray-matter of Mindboggle ^11^. Volume-based spatial normalization to two standard spaces (MNI152NLin2009cAsym, MNI152NLin6Asym) was performed through nonlinear registration with antsRegistration (ANTs 2.5.0), using brain-extracted versions of both T1w reference and the T1w template. The following templates were selected for spatial normalization and accessed with TemplateFlow (23.1.0)^12^: ICBM 152 Nonlinear Asymmetrical template version 2009c ^13^ (TemplateFlow ID: MNI152NLin2009cAsym), FSL’s MNI ICBM 152 non-linear 6th Generation Asymmetric Average Brain Stereotaxic Registration Model ^14^ (TemplateFlow ID: MNI152NLin6Asym). Grayordinate “dscalar” files containing 91k samples were resampled onto fsLR using the Connectome Workbench ^15^.

*Functional data preprocessing*

For each of the BOLD runs found per subject (across all tasks and sessions), the following preprocessing was performed. First, a reference volume was generated, using a custom methodology of fMRIPrep, for use in head motion correction. Head-motion parameters with respect to the BOLD reference (transformation matrices, and six corresponding rotation and translation parameters) are estimated before any spatiotemporal filtering using mcflirt ^16^. Where appropriate (Melbourne and Brisbane datasets), the estimated fieldmap was then aligned with rigid-registration to the target EPI (echo-planar imaging) reference run. The field coefficients were mapped on to the reference EPI using the transform. The BOLD reference was then co-registered to the T1w reference using bbregister (FreeSurfer) which implements boundary-based registration ^17^. Co-registration was configured with six degrees of freedom. Several confounding time-series were calculated based on the preprocessed BOLD: framewise displacement (FD), DVARS and three region-wise global signals. FD was computed using two formulations following Power (absolute sum of relative motions), and Jenkinson (relative root mean square displacement between affines). FD and DVARS are calculated for each functional run, both using their implementations in Nipype ^18,19^. The three global signals are extracted within the CSF, the WM, and the whole-brain masks. The confound time series derived from head motion estimates and global signals were expanded with the inclusion of temporal derivatives and quadratic terms for each ^20^. Frames that exceeded a threshold of 0.5 mm FD or 1.5 standardized DVARS were annotated as motion outliers. Additional nuisance timeseries are calculated by means of principal components analysis of the signal found within a thin band (crown) of voxels around the edge of the brain, as proposed by ^21^. The BOLD time-series were resampled onto the left/right-symmetric template “fsLR” using the Connectome Workbench ^15^. Grayordinates files containing 91k samples were also generated with surface data transformed directly to fsLR space and subcortical data transformed to 2 mm resolution MNI152NLin6Asym space. All resamplings can be performed with a single interpolation step by composing all the pertinent transformations (i.e., head-motion transform matrices, susceptibility distortion correction when available, and co-registrations to anatomical and output spaces). Gridded (volumetric) resamplings were performed using nitransforms, configured with cubic B-spline interpolation.

Many internal operations of fMRIPrep use Nilearn 0.10.2 ^22^, mostly within the functional processing workflow. For more details of the pipeline, see the section corresponding to workflows in fMRIPrep’s documentation.

#### Cortico-striatal circuit mapping

To define connections of interest we used an independent dataset of 250 unrelated participants from the Human Connectome Project (HCP) to map the four circuits of interest. To do so, four seed-to-region analyses were performed using a high-resolution subcortical brain atlas ^23^. A one-sample *t*-test was performed and the 10 highest (“most functionally connected”) RSFC edges were considered for each hemisphere in each of the four seed regions (Nucelus Accumbens (NAcc), dorsal Caudate (dCaud), dorsal Putamen (dPut), and ventral Putamen (vPut)).

It’s important to note that the subcortical regions in the brain parcellation required for meta-matching were larger than the high-resolution mapping template that we adopted for mapping the circuits of interest (**Supplementary Figure 1**). Thus, when deriving the RSFC feature weights for the circuits of interest the closest subcortical region was chosen (in addition to the exact same cortical regions). This means that, while the circuits were derived from a highly specific *dorsal* caudate region, when averaging the RSFC feature weights the more lenient entire caudate region was used.

Supplementary Tables

**Supplementary Table 1. Summary of RSFC feature weights across functional networks**

| Network | Median FW^1e-3^ (IQR) | Percentile | *p*_FDR_ |
| --- | --- | --- | --- |
| Control / frontoparietal (FPN) | -0.74 (-1.07 - -0.39) |  |  |
|  | -1.96 (-2.32 - -1.67) | 0.035 | 0.046 |
| Default-mode network (DMN) | 0.36 (-0.04 - 0.77) |  |  |
|  | -1.26 (-1.46 - -1.04) | 0.008 | 0.034 |
| Dorsal Attention (DAt) | -1.15 (-1.5 - -0.79) |  |  |
|  | -1.67 (-2.09 - -1.22) | 0.934 | 0.066 |
| Limbic (Lim) | 0.22 (-0.08 - 0.47) |  |  |
|  | 1.33 (0.9 - 1.8) | 0.004 | 0.034 |
| Salience / Ventral Attention (VAt) | -0.74 (-1.07 - -0.39) |  |  |
|  | -1.96 (-2.32 - -1.67) | 0.025 | 0.04 |
| Somatomotor (SM) | 0.36 (-0.04 - 0.77) |  |  |
|  | -1.26 (-1.46 - -1.04) | 0.014 | 0.034 |
| Subcortical (SC) | -1.15 (-1.5 - -0.79) |  |  |
|  | -1.67 (-2.09 - -1.22) | 0.94 | 0.066 |
| Visual (Vis) | 0.22 (-0.08 - 0.47) |  |  |
|  | 1.33 (0.9 - 1.8) | 0.983 | 0.034 |

FW; feature weights, IQR; interquartile range.

**Supplementary Table 2. Summary of predictive weights in circuits of interest**

| Pathway | Median RSFC FW^1e-3^ (IQR) | Percentile compared to Null Model | |
| --- | --- | --- | --- |
|  |  | *Permutation* | *Subcortical –*  *Cortical* |
| NAcc | 0.20 (-0.37 - 0.74) | 0.9226 | 0.4836 |
| dCaud | -2.77 (-3.55 - -1.96) | 0.0124 | 0.0216 |
| dPut | -2.33 (-2.93 - -1.71) | 0.0076 | 0.039 |
| vPut | -1.96 (-2.57 - -1.41) | 0.0098 | 0.0668 |

FW; feature weights, IQR; interquartile range, NAcc; Nucelus Accumbens, dCaud; dorsal Caudate, dPut; dorsal Putamen, vPut; ventral Putamen.

**Supplementary Table 3. Top 50 Meta-matching component predictive feature weights**

| Source Dataset | Method | Meta-matching component | Median FW^1e-3^ (IQR) | Percentile | *p*_FDR_ |
| --- | --- | --- | --- | --- | --- |
| HBN | RSFC KRR | NIH7_Card | -23.56 (-25.82 - -20.6) | 0 | <0.001 |
| HBN | RSFC KRR | NIH7_Flanker | -25.11 (-27.93 - -22.05) | 0 | <0.001 |
| eNKI | RSFC KRR | DKEFSCWI_14 | 12.55 (11.47 - 13.67) | 0.999 | 0.055 |
| HBN | RSFC KRR | SDQ_Peer_Problems | 14.33 (12.65 - 16.11) | 0.999 | 0.055 |
| GSP | RSFC KRR | TCI_HarmAvoidance | 17.38 (15.84 - 18.88) | 1 | 0.055 |
| UKBB | RSFC KRR | BP_eye_C5 | 27.39 (23.9 - 30.57) | 0.998 | 0.059 |
| UKBB | RSFC KRR | Digit-o_C6 | -6.09 (-6.77 - -5.31) | 0.002 | 0.059 |
| eNKI | RSFC KRR | VF_41 | 12.1 (10.76 - 13.43) | 0.999 | 0.059 |
| eNKI | RSFC KRR | WIAT_06 | 4.82 (4.32 - 5.31) | 0.998 | 0.059 |
| HBN | RSFC KRR | SDQ_Prosocial | -7.17 (-8.42 - -6.0) | 0.002 | 0.059 |
| UKBB | RSFC KRR | Travel | -8.93 (-10.24 - -7.73) | 0.002 | 0.059 |
| eNKI | RSFC KRR | DKEFSCWI_18 | 11.11 (9.9 - 12.34) | 0.999 | 0.059 |
| UKBB | RSFC KRR | Sleep | 9.78 (8.6 - 11.15) | 0.998 | 0.059 |
| GSP | RSFC KRR | BISBAS_BAS_Drive | -3.73 (-4.17 - -3.31) | 0.001 | 0.059 |
| ABCD | RSFC KRR | cbcl_scr_syn_somatic_r | -4.65 (-5.33 - -3.82) | 0.003 | 0.061 |
| ABCD | RSFC KRR | cbcl_scr_syn_social_r | -12.18 (-14.24 - -10.13) | 0.008 | 0.061 |
| HBN | RSFC KRR | ARI_S_Total_Score | -15.42 (-17.84 - -13.14) | 0.005 | 0.061 |
| ABCD | RSFC KRR | cbcl_scr_syn_aggressive_r | -16.31 (-18.63 - -14.02) | 0.004 | 0.061 |
| eNKI | RSFC KRR | VF_37 | 1.75 (1.37 - 2.09) | 0.993 | 0.061 |
| HBN | RSFC KRR | NIH7_Pattern | -16.2 (-18.19 - -14.12) | 0.004 | 0.061 |
| eNKI | RSFC KRR | VF_54 | 4.13 (3.33 - 4.94) | 0.993 | 0.061 |
| ABCD | RSFC RSFC | cbcl_scr_syn_rulebreak_r | -17.91 (-20.51 - -15.3) | 0.004 | 0.061 |
| HBN | DNN KRR | SRS_MOT_T | -197.19 (-226.1 - -162.57) | 0.007 | 0.061 |
| UKBB | RSFC KRR | Alcohol_1 | -4.0 (-4.74 - -3.14) | 0.006 | 0.061 |
| UKBB | RSFC KRR | Body_C3 | 29.99 (25.83 - 34.27) | 0.995 | 0.061 |
| eNKI | RSFC KRR | DKEFSTMT_18 | 4.7 (3.98 - 5.34) | 0.998 | 0.061 |
| ABCD | RSFC KRR | upps_y_ss_negative_urgency | -10.72 (-11.97 - -9.46) | 0.005 | 0.061 |
| UKBB | RSFC KRR | Illness_C4 | -11.6 (-13.71 - -9.87) | 0.008 | 0.061 |
| eNKI | RSFC KRR | DKEFSTMT_20 | 8.12 (6.6 - 9.52) | 0.998 | 0.061 |
| UKBB | RSFC KRR | Age_edu | -4.61 (-5.57 - -3.65) | 0.007 | 0.061 |
| UKBB | RSFC KRR | Work | -24.61 (-27.36 - -22.02) | 0.004 | 0.061 |
| UKBB | RSFC KRR | Neuro | 13.16 (11.36 - 14.96) | 0.996 | 0.061 |
| HBN | RSFC KRR | ASSQ_Total | 13.64 (11.11 - 16.01) | 0.997 | 0.061 |
| eNKI | RSFC KRR | DKEFSTMT_28 | 3.61 (3.09 - 4.1) | 0.994 | 0.061 |
| eNKI | RSFC KRR | TOWER_53 | -16.32 (-18.19 - -14.24) | 0.004 | 0.061 |
| eNKI | RSFC KRR | PTSDCH_55 | -6.92 (-7.96 - -5.9) | 0.007 | 0.061 |
| UKBB | RSFC KRR | BP_eye_C4 | -13.45 (-15.9 - -10.81) | 0.007 | 0.061 |
| ABCD | RSFC KRR | upps_y_ss_positive_urgency | -15.31 (-17.46 - -13.15) | 0.006 | 0.061 |
| eNKI | RSFC KRR | VF_49 | 6.08 (5.03 - 7.13) | 0.995 | 0.061 |
| UKBB | RSFC KRR | Genetic_C1 | -43.63 (-53.7 - -35.11) | 0.007 | 0.061 |
| GSP | RSFC KRR | DOSPERT_perception | -8.74 (-10.58 - -6.76) | 0.006 | 0.061 |
| GSP | RSFC KRR | TCI_RewardDependence | -9.43 (-11.28 - -7.52) | 0.006 | 0.061 |
| GSP | RSFC KRR | BISBAS_BIS | 9.1 (7.83 - 10.63) | 0.996 | 0.061 |
| eNKI | RSFC KRR | DKEFSCWI_40 | 10.22 (8.5 - 11.89) | 0.993 | 0.061 |
| HBN | RSFC KRR | SRS_COG_T | 13.36 (11.3 - 15.29) | 0.996 | 0.061 |
| GSP | RSFC KRR | NEO_E | -9.06 (-10.67 - -7.51) | 0.006 | 0.061 |
| HBN | RSFC KRR | ARI_P_Total_Score | -6.92 (-8.3 - -5.63) | 0.005 | 0.061 |
| GSP | RSFC KRR | NEO_N | 6.33 (5.25 - 7.37) | 0.994 | 0.061 |
| HBN | RSFC KRR | SRS_MOT_T | -21.49 (-25.34 - -17.68) | 0.007 | 0.061 |
| eNKI | RSFC KRR | DKEFSCWI_20 | 2.58 (2.13 - 3.04) | 0.996 | 0.061 |

Note: Method refers to process by which the phenotype prediction was made. RSFC KRR denotes the kernel ridge regression models, whereas RSFC DNN refers to using the deep neural network as input features. Further information about the phenotypes used are available in the original multilayer meta-matching manuscript ^24^**.** FW; feature weights, IQR; interquartile range.

**Supplementary Table 4. Relationship between regional gene expression and RSFC brain connectivity features**

| Model | Location | Gene | *r* | *p* |
| --- | --- | --- | --- | --- |
| Meta-match | Subcortex | ADCK1 | 0.16 | 0.251 |
|  |  | GRID2 | 0.39 | 0.07 |
|  |  | KIT | 0.27 | 0.145 |
|  |  | WDR7 | -0.64 | 0.005 |
|  | Cortical surface | ADCK1 | -0.14 | 0.025 |
|  |  | GRID2 | -0.31 | < 0.001 |
|  |  | KIT | 0.12 | 0.059 |
|  |  | WDR7 | -0.3 | < 0.001 |

Note: the reported *p* values were FDR corrected across comparisons (8 per model) after the performing the spin test.

Supplementary Figures

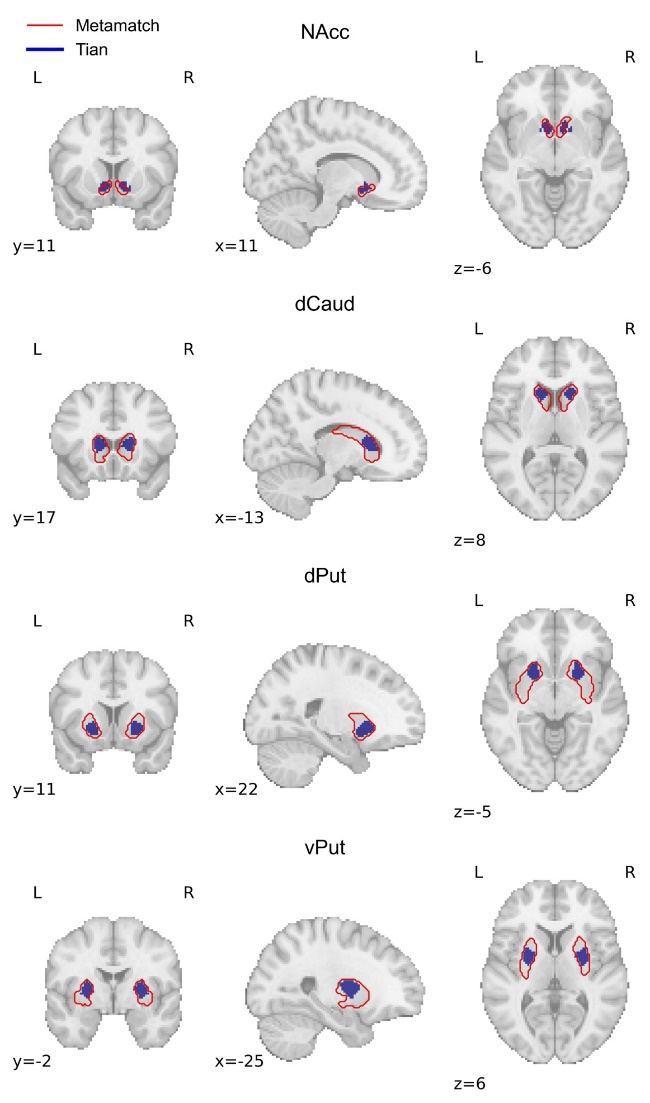

**Supplementary Figure 1. Comparison between subcortical seed regions in the high-resolution Tian atlas (blue, used in the HCP pathway mapping) ^23^ and the Meta-matching atlas (red, used in the classification analyses).**

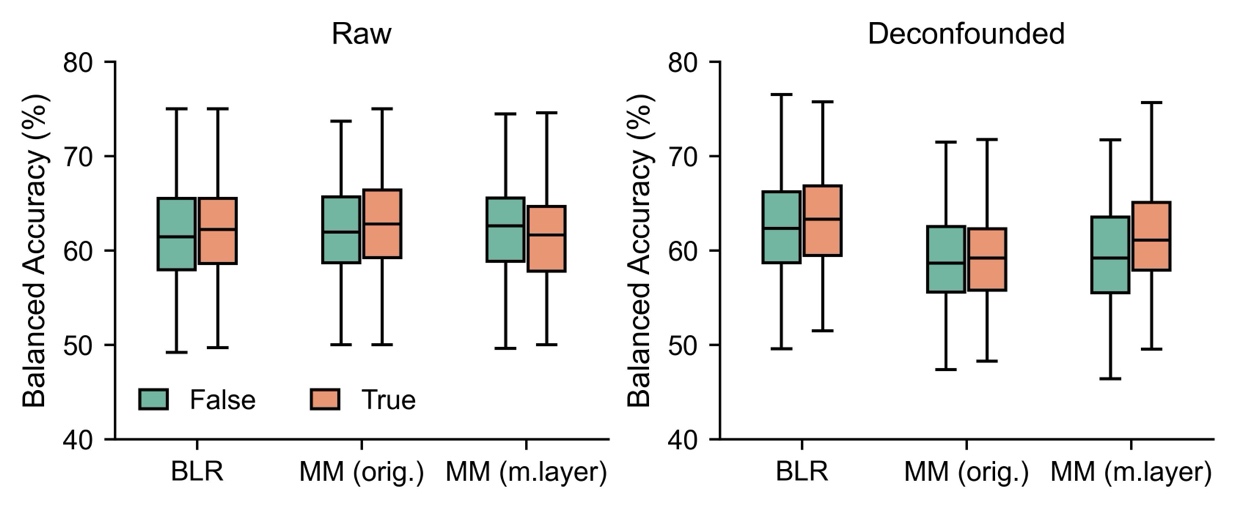

**Supplementary Figure 2. Classification performance across models, denoising and deconfounding approaches.** The orange (True) and green (False) denote the presence, or lack thereof global signal regression in the denoising pipeline. The left panel shows balanced accuracy (y-axis) for each model in the raw data prior to deconfounding. The right panel shows the results post-deconfounding. BLR; baseline logistic regression, MM (orig.) refers to the original meta-matching model ^25^ and MM (m.layer) refers to the multilayer meta-matching model ^24^ which was used in the main analyses. Boxplots: centre line; median; box limits; upper and lower quartiles; whiskers; 1.5x interquartile range.
